# In-home validation of wrist Actigraphy against portable electroencephalography for sleep assessment in older adults

**DOI:** 10.64898/2026.01.15.26344168

**Authors:** Naoki Deguchi, Sho Hatanaka, Kaori Daimaru, Kazushi Maruo, Hiroyuki Sasai

## Abstract

**Background:** While accurate sleep measurement is vital for older adults, the validity of actigraphy (AG) in free-living environments remains controversial, particularly given the flexible sleep-wake schedules common in this demographic. To address this uncertainty, we assessed the accuracy of wrist AG against in-home portable electroencephalography (EEG) among community-dwelling older adults.

**Methods:** Community-dwelling older adults underwent concurrent sleep monitoring using a portable EEG device and a wrist-worn AG for five consecutive nights whenever possible, with monitoring extended to up to seven nights when feasible. Key sleep parameters, including total sleep time (TST), sleep onset latency (SOL), wake after sleep onset (WASO), and sleep efficiency, were derived from both devices. Measurement agreement was assessed using Bland–Altman plots and multilevel modeling, while reliability and accuracy were quantified via intraclass correlation coefficients (ICCs) and mean absolute percentage error (MAPE).

**Results:** Forty-nine adults contributed 217 nights of recordings. On average, AG slightly overestimated TST and sleep efficiency and underestimated SOL and WASO compared with EEG. Single-measure ICCs were 0.73 for TST and 0.38 for WASO (0.84 and 0.55 for averages across nights), and the MAPE was 11% for TST but exceeded 50% for SOL and WASO, indicating poor accuracy for these indices.

**Conclusion:** In community-dwelling older adults, wrist AG yielded acceptably accurate estimates of average TST, supporting its use in epidemiological monitoring of sleep duration. However, large errors for SOL and WASO indicate that portable EEG- or polysomnography-based assessment remains indispensable when precise evaluation of sleep initiation and nocturnal wakefulness is required.

## 1. Introduction

Sleep is essential for maintaining numerous physiological functions, including brain activity, metabolism, appetite regulation, immune responses, hormonal balance, and cardiovascular health [1,2]. In older adults, reductions in slow-wave sleep, increased nighttime awakenings, and earlier sleep onset are linked to a higher prevalence of sleep disorders, including insomnia and sleep apnea [3]. The increasing prevalence of sleep disorders and difficulty initiating sleep among older adults impacts their health and quality of life [4]. Conversely, a potential discrepancy has been suggested between subjective sleep assessments based on self-reports and objective sleep indicators [5]. Therefore, accurate and objective sleep assessment methods for older adults are crucial for promoting their well-being.

Polysomnography (PSG) is the gold standard for sleep assessment; however, its dependence on high-cost and sophisticated equipment and trained professionals limit its feasibility in natural settings [6,7]. To overcome these limitations, portable electroencephalography (EEG) devices have emerged as a viable alternative, enabling multi-night monitoring at home [8–10]. These devices offer sleep-stage detection accuracy comparable to PSG across diverse populations [10] and allow for extended monitoring with minimal disruption to daily routines. Moreover, portable EEG devices support long-term sleep monitoring in real-world conditions, accounting for variations in work schedules, physical activity, and sleep habits. Recent validation studies and systematic reviews have shown that portable EEG provides sleep indices that closely track in-laboratory PSG in both clinical and community samples, supporting its role as a realistic “second-best” or pragmatic reference standard for free-living sleep assessment [8–10]. Nevertheless, despite reducing infrastructure requirements and participant burden, portable EEG still necessitates electrode placement and technical maintenance—factors that constrain its scalability in large-scale epidemiological cohorts and routine geriatric care.

Actigraphy (AG), by contrast, is cost-effective for measuring key sleep parameters (e.g., total sleep time [TST], sleep onset latency [SOL], wake after sleep onset [WASO], and sleep efficiency) and imposes minimal burden on participants, allowing for long-term data collection in naturalistic settings [11]. It is widely utilized in large-scale epidemiological studies [12,13]. While AG shows moderate-to-strong concordance with PSG in general populations, its validity remains under-examined in older adults, who often exhibit fragmented sleep architectures, including increased WASO and reduced TST [14–16]. Although portable EEG devices allow detailed at-home sleep assessment, studies validating AG against portable EEG in older adults are strikingly limited. Given the unique sleep profiles of older adults, rigorous validation in this population is essential to ensure accurate interpretation of AG-derived sleep metrics. Accordingly, AG could be particularly useful for continuous and unobtrusive sleep monitoring in older adults, potentially improving real-world assessments and bridging the gap between clinical evaluations and daily life.

This study aimed to assess the free-living validity of wrist AG as an objective sleep assessment tool in community-dwelling older adults, using a portable EEG device as a pragmatic reference standard that retains much of PSG’s physiological fidelity while enabling in-home recordings. To our knowledge, this is the first study to validate multi-night wrist AG directly against in-home portable EEG in community-dwelling older adults without major sleep disorders. By quantifying the accuracy of TST, SOL, WASO, and sleep efficiency under real-life conditions, we sought to clarify which AG indices are suitable for large-scale population monitoring of sleep duration and fragmentation and which still require EEG- or PSG-based assessment.

## 2. Methods

### 2.1 Design and settings

This study was conducted as an ancillary investigation within the Smart Watch Innovation for Next Geriatrics & Gerontology (SWING-Japan) project [17–19], an initiative designed to enhance health outcomes in older adults through the integration of wearable-derived physical activity and sleep data. Participants for this validation study were recruited between May and June 2023 from the Itabashi Longitudinal Study on Aging (Itabashi LSA) [20, 21], a constituent cohort of SWING-Japan. The Itabashi LSA is an ongoing longitudinal study of community-dwelling individuals aged 70–85 residing in Itabashi Ward, Tokyo. Candidates for the current sleep health checkup were identified based on baseline assessments performed during the February 2023 survey wave.

### 2.2 Participants Participants and Eligibility

Eligible participants were community-dwelling adults aged 70–85 years from the Itabashi LSA cohort. To ensure a sample representative of healthy aging without significant sleep pathology, individuals were excluded based on the following criteria: (1) a clinical diagnosis of insomnia (ICSD-3); (2) suspected sleep disorders (Pittsburgh Sleep Quality Index > 6 [22]); (3) habitual sleep duration outside the 6–9 hour range; (4) a body mass index < 18.5 or ≥ 30.0 kg/m²; (5) lower global cognitive function (Mini-Mental State Examination [MMSE] score < 28 [23]); (6) clinical depression or significant depressive symptoms ((Geriatric Depression Scale–15 [GDS-15]score > 10 [24]); or (7) current smoking.

#### Recruitment and Screening

Potential candidates were identified through the Itabashi LSA database and mailed a study overview alongside a secondary screening questionnaire. This questionnaire assessed logistical eligibility, including: (1) ability to tolerate wrist-worn and portable EEG devices for five consecutive nights (up to seven nights when feasible); (2) use of sleep medications (prescription or over-the-counter) within the past month; (3) willingness to refrain from caffeine within five hours of bedtime and alcohol throughout the study; (4) presence of bed-sharing with individuals or animals; (5) planned overnight stays away from home; and (6) recent or scheduled night-shift work. Individuals meeting all criteria were invited to an orientation session for detailed briefing and formal enrollment.

#### Ethics and Compensation

Written informed consent was obtained from all participants prior to data collection. As compensation, participants received a 10,000-yen gift card upon completion, regardless of data yield, to mitigate any undue influence on participant behavior. The study protocol was approved by the Research Ethics Committee of the Tokyo Metropolitan Institute for Geriatrics and Gerontology (Approval No.: R022-099) and adhered to the principles of the Declaration of Helsinki.

### 2.3 Baseline Characteristics

Demographic and clinical characteristics were obtained from self-administered questionnaires completed as part of the Itabashi LSA baseline assessment. Participants self-reported their age, sex, living arrangement, employment status, subjective health, and Pittsburgh Sleep Quality Index [22], Mini-Mental State Examination score [23], and Geriatric Depression Scale score [24].

### 2.4 Portable EEG Device (reference)

Sleep variables were recorded using the portable EEG device Insomnograf K2 (S’UIMIN Inc., Tokyo, Japan) (Figure 1A). This lightweight device (162 g) employs flexible adhesive electrodes that are easy to attach and remove, making it particularly suitable for older adults. The device’s concordance with conventional PSG has been validated, with a κ coefficient of 0.71 for the classification of five sleep stages: Wake, rapid eye movement sleep (REM), stage 1 of non-REM sleep (NREM1), stage 2 of non-REM sleep (NREM2), and stage 3 of non-REM sleep (NREM3) [25].

**Figure 1.**
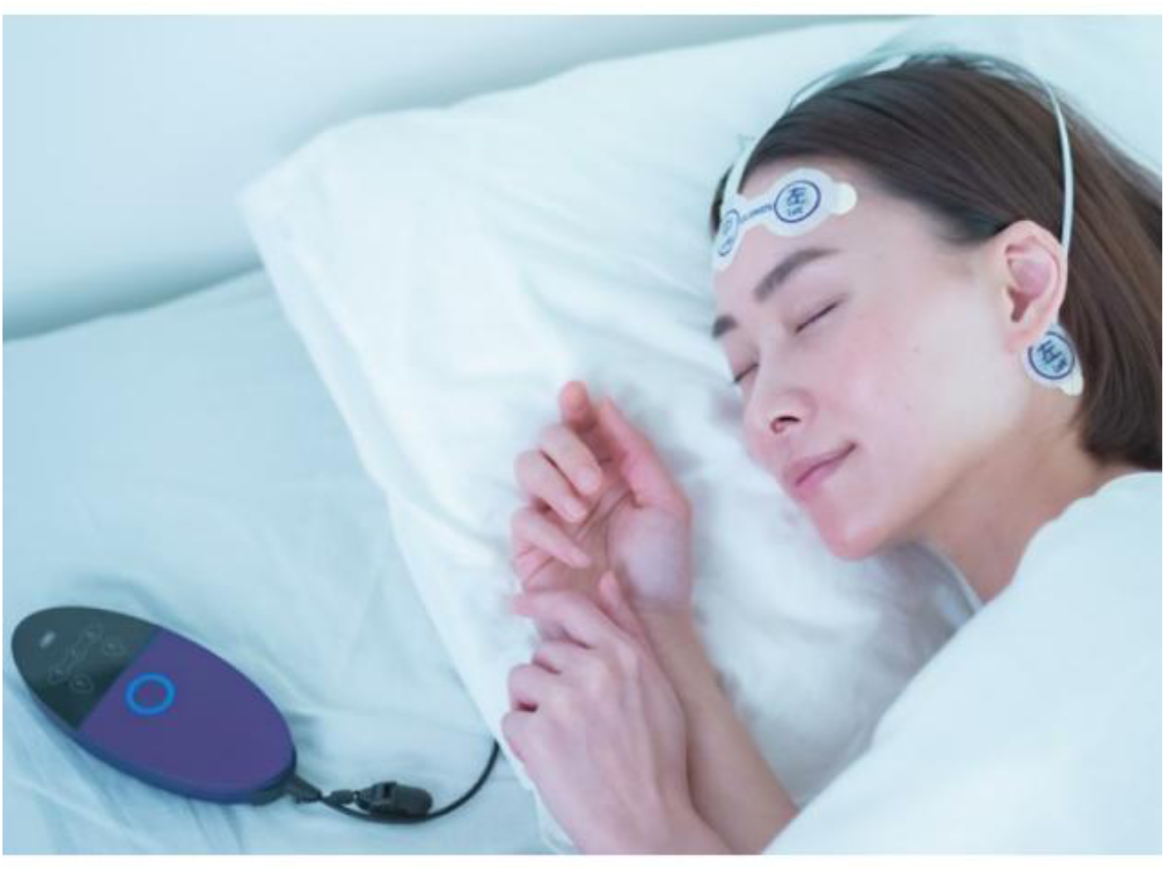
Images of (A) InSomnograf K2®* (S’UIMIN Inc., Japan) *Publicly available at https://www.suimin.co.jp/

The recording system follows the international 10–20 system and includes four EEG electrodes (Fp1, Fp2, M1, and M2) and one reference electrode (Fpz). Sleep-stage analysis utilized four EEG derivations (Fp1–M2, Fp2–M1, Fp1–Mean M, Fp2–Mean M), with Fp1–Fp2 and Fp2–Fp1 serving as left and right electrooculograms, respectively, and M1–M2 as the submental electromyogram. Accordingly, portable EEG was used as the pragmatic reference device for evaluating AG-derived sleep indices in this study [18,25].

### 2.5 Actigraphy

Participants wore an ActiGraph wGT3X-BT+ (Ametris, Pensacola, FL, USA) on the non-dominant wrist during all nights when the portable EEG device was used. This device, measuring 46 mm × 33 mm × 15 mm and weighing 19 g, is equipped with a triaxial accelerometer and collects data at 80 Hz using a 12-bit analog-to-digital converter within a range of ±6 g.

Raw acceleration data were downloaded from the device’s non-volatile memory using ActiLife software (version 6.13.4; ActiGraph LLC). Within ActiLife, the 80-Hz data were reintegrated into 60-s epochs and classified as “sleep” or “wake” using the Cole–Kripke algorithm [26,27]. The resulting 60-s epoch-level classifications were then exported and further processed in R (version 4.2.2; R Foundation for Statistical Computing, Vienna, Austria) for the calculation of sleep indices, as described in Section 2.7.

### 2.6 Data collection procedure

Based on prior studies suggesting that multi-night monitoring improves reliability and reduces measurement error [28], participants were asked to wear both the portable EEG device and the ActiGraph for five consecutive nights whenever possible. five consecutive nights whenever possible. When feasible, monitoring was extended to a maximum of seven nights to increase the number of valid recordings. For feasibility, participants who provided at least three nights with valid data from both devices were included in the analysis. During the monitoring period, participants recorded “lights-off time” and “wake-up time” each night in designated sleep diaries to allow calculation of time in bed (TIB).

Each evening, participants were instructed to attach the portable EEG device electrodes after bathing or showering, press the record button immediately before going to bed to start the recording, and press the stop button upon final awakening. The ActiGraph wGT3X-BT+ was worn on the non-dominant wrist during all nights when the portable EEG device was used. Participants were instructed to put on the ActiGraph before going to bed and to keep it on until they woke up in the morning.

Both devices were distributed after participants received in-person instructions on their use at the time of enrollment. After completing the monitoring period, participants returned all devices to the research team.

### 2.7 Data Processing

Data were synchronized across the portable EEG and ActiGraph data based on their respective epoch-level classifications, using the first 60-s epoch. For all devices, scoring was conducted independently and blinded to other results. An independent researcher subsequently imported the epoch-level data into R (version 4.2.2) and aggregated them to calculate the following standardized sleep indices: (a) TST: total minutes scored as “sleep” within TIB; (b) SOL: time from lights-off to sleep onset; (c) WASO: time scored as “wake” from sleep onset to final awakening; and (d) sleep efficiency (%) = TST / TIB×100. SOL was defined as the time from “lights-off” to the first epoch classified as “sleep,” where “sleep” was operationalized as the absence of any “wake” epochs for at least one consecutive minute [29,30].

For the portable EEG, a sleep EEG analyst with over 5 years of experience scored each 60-s epoch into five states: wake, REM, NREM1, NREM2, and NREM3. Epochs classified as REM or any non-REM stage were aggregated and treated as “sleep,” while wake epochs were treated as “wake.” Scoring was conducted blinded to other results, and the datasets were subsequently linked by an independent researcher.

For the ActiGraph wGT3X-BT+, the collected acceleration data were analyzed using the ActiLife software (version 6.13.4; ActiGraph LLC) with the manufacturer-implemented Cole–Kripke algorithm [26,27]. Within ActiLife, the 80-Hz raw signals were reintegrated into 60-s epochs and classified as either “wake” or “sleep” based on accelerometer-derived activity counts. The resulting epoch-level data are expressed as binary states (wake/sleep), and some computational details, such as the internal thresholds and device operating parameters, are not publicly disclosed. Sleep indices were derived from these 60-s epoch classifications using the same definitions as those applied to the portable EEG.

### 2.8. Statistical Analysis General procedures

Continuous variables were summarized as mean ± standard deviation, and categorical variables as frequencies (percentages). Nights with more than 5% missing data for any variable were excluded, and participants with at least three valid nights were retained. All analyses accounted for the hierarchical structure (up to seven nights per participant). Analyses were performed using SPSS version 29.0 (IBM Corp., Armonk, NY, USA) and R version 4.2.2 (R Foundation for Statistical Computing, Vienna, Austria). Statistical significance was set at a two-sided P value < 0.05. To address the study aims, the following analyses were conducted.

#### Participant characteristics and portable EEG sleep parameters

Participant characteristics were summarized descriptively. Overall sleep parameters based on portable EEG were summarized using an intercept-only multilevel model to estimate the overall mean and standard error of each sleep parameter (TST, SOL, WASO, and sleep efficiency) across all participants, accounting for repeated nightly observations within individuals [31].

#### Systematic bias and limits of agreement

To examine systematic bias and 95% limits of agreement (LoA) between AG and portable EEG, Bland–Altman plots were constructed with the X-axis representing the average of the two devices and the Y-axis representing their paired differences (AG minus portable EEG). To account for repeated nightly measurements within individuals, multilevel Bland–Altman models were fitted to estimate the overall mean difference and 95% LoA [32,33]. Fixed and proportional bias were examined by including the average of the two devices as a fixed effect and testing the intercept (fixed bias) and slope (proportional bias). Because SOL and WASO were markedly skewed, these variables were log-transformed prior to Bland–Altman plotting and multilevel modeling.

#### Magnitude of disagreement

To quantify the magnitude of absolute and relative disagreement between AG and portable EEG, mean absolute error (MAE) and mean absolute percentage error (MAPE) were calculated for each sleep parameter. MAE was defined as the mean of |AGᵢ − portable EEGᵢ| across all observations, and MAPE as the mean of (|AGᵢ − portable EEGᵢ| / |portable EEGᵢ| × 100) across all observations.

#### Reliability between devices

To evaluate agreement consistency between AG and portable EEG, ICCs were computed using a two-way mixed-effects model with absolute agreement for both single and average measures [34, 35]. ICC values were interpreted as “excellent” (≥0.75), “fair to good” (0.40–0.75), and “poor” (<0.40) [36].

## 3. Results

### 3.1 Participants

Initial invitations were sent to 175 individuals from the Itabashi LSA participants. Of these, 72 expressed a willingness to participate, and 61 provided written informed consent (Figure 2). The final analysis comprised 49 participants who completed simultaneous measurements with the portable EEG and AG devices, providing data for 217 nights.

**Figure 2.**
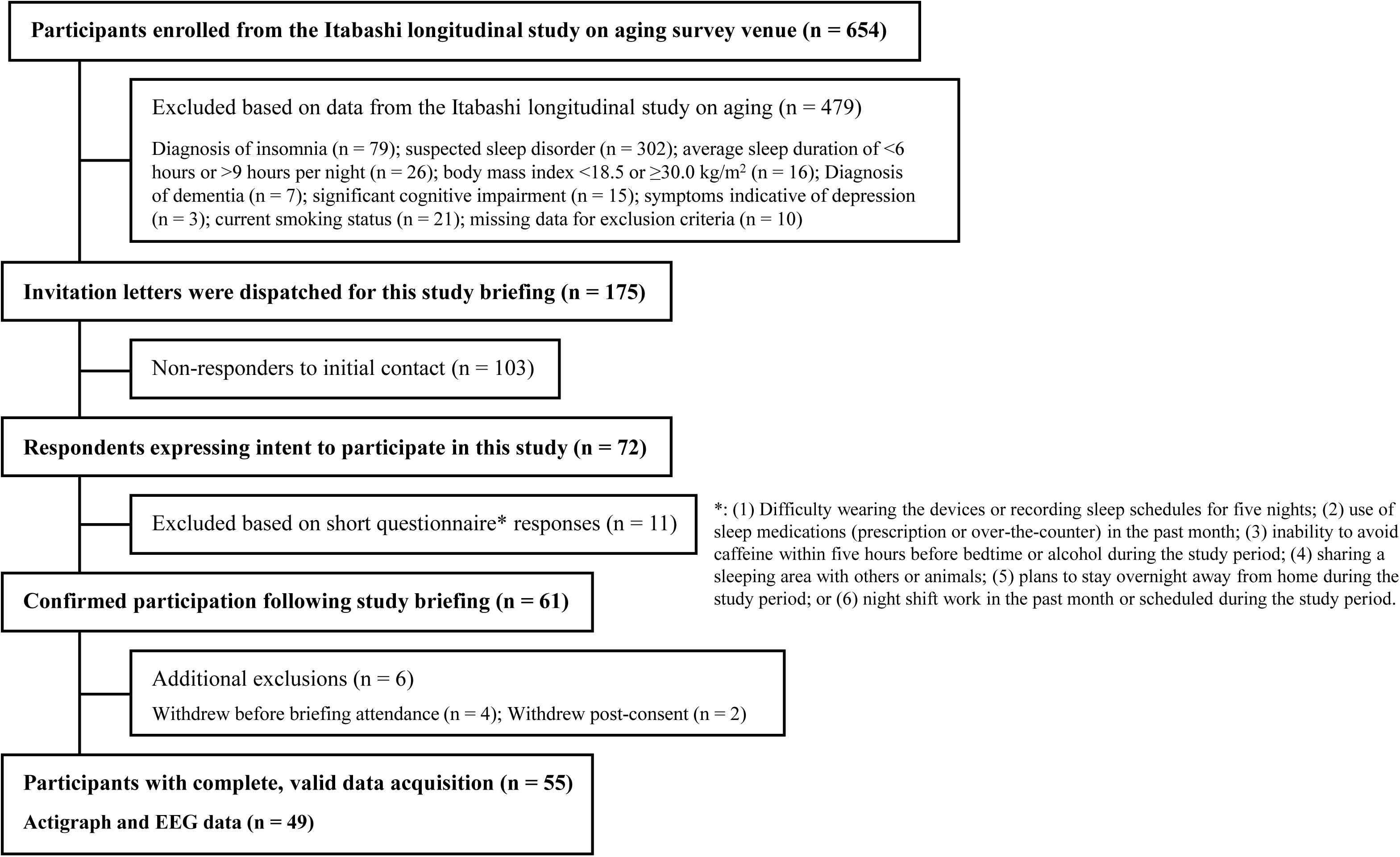
Participant recruitment flow chart

### 3.2 Participant characteristics and portable EEG sleep parameters

Participant characteristics are summarized in Table 1. The mean age was 73.9 ± 3.4 years (range, 70–83 years), and 61.2% were men. Based on the portable EEG, the overall mean nightly sleep parameters were 345 min for TST, 13.3 min for SOL, 57.2 min for WASO, and 81.2% for sleep efficiency (Table 2).

**Table 1.** Participant characteristics.

|  |  |
| --- | --- |
| Demographics |  |
| **Age, years** | 73.9 ± 3.4 |
| **Sex, male, n (%)** | 30 (61.2) |
| **Body mass index, kg/m<sup>2</sup>** | 23.3 ± 2.9 |
| **Living alone, n (%)** | 8 (16.3) |
| **Educational attainment, n (%)** |  |
| No formal education, elementary or junior high school | 0 (0) |
| High school, junior college, or vocational school | 24 (49.0) |
| University or graduate school | 25 (51.0) |
| **Employment status** |  |
| Work full-time | 2 (4.1) |
| Work part-time / irregular | 19 (38.8) |
| Not working | 28 (57.1) |
| **Subjective health, n (%)** |  |
| Excellent | 13 (26.5) |
| Very good | 20 (40.8) |
| Good | 14 (28.6) |
| Fair | 1 (2.0) |
| Poor | 0 (0) |
| Missing data | 1 (2.0) |
| **Body pain, yes, n (%)** | 19 (38.8) |
| Clinical Assessments |  |
| **Pittsburgh Sleep Quality Index, points** | 3.2 ± 1.1 |
| **Geriatric Depression Scale, points** | 3.3 ± 2.8 |
| **Mini-Mental State Examination, points** | 29.2 ± 0.9 |

*Data are presented as mean ± standard deviation.*

**Table 2.**
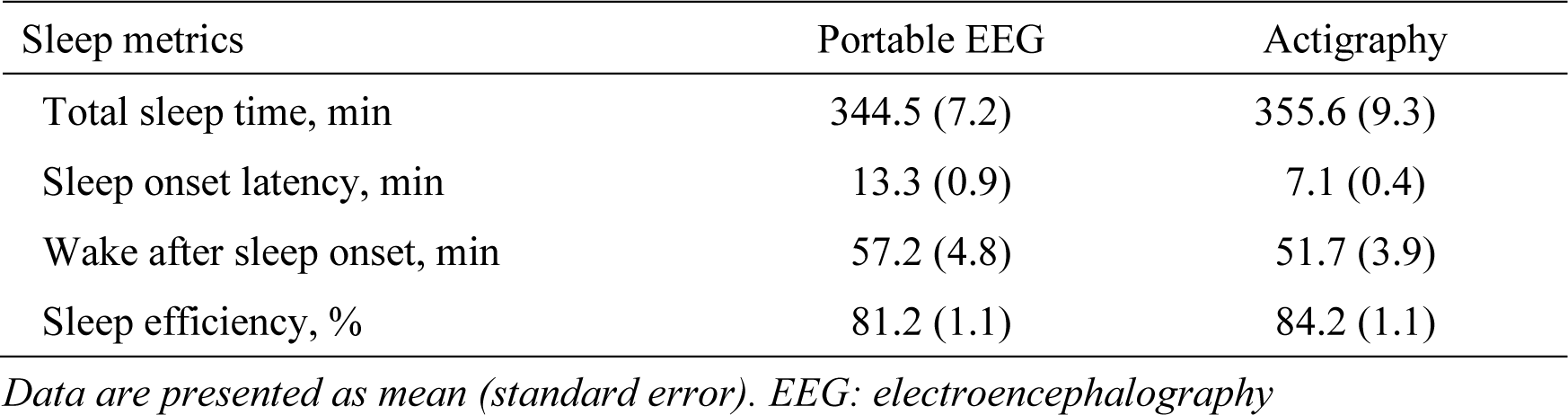
Descriptive sleep metrics for portable EEG and actigraphy.

### 3.3 Systematic bias and limits of agreement

BA analysis indicated systematic differences between the two devices (Figure 3). Compared with portable EEG, AG overestimated TST by a mean of 10.4 min (LoA: −78 to 98) and underestimated SOL by 5.8 min (LoA: −22 to 10). AG underestimated WASO by 1.6 min on average (LoA: −82 to 79) and overestimated sleep efficiency by 2.2 percentage points (LoA: −18.4 to 22.9).

**Figure 3.**
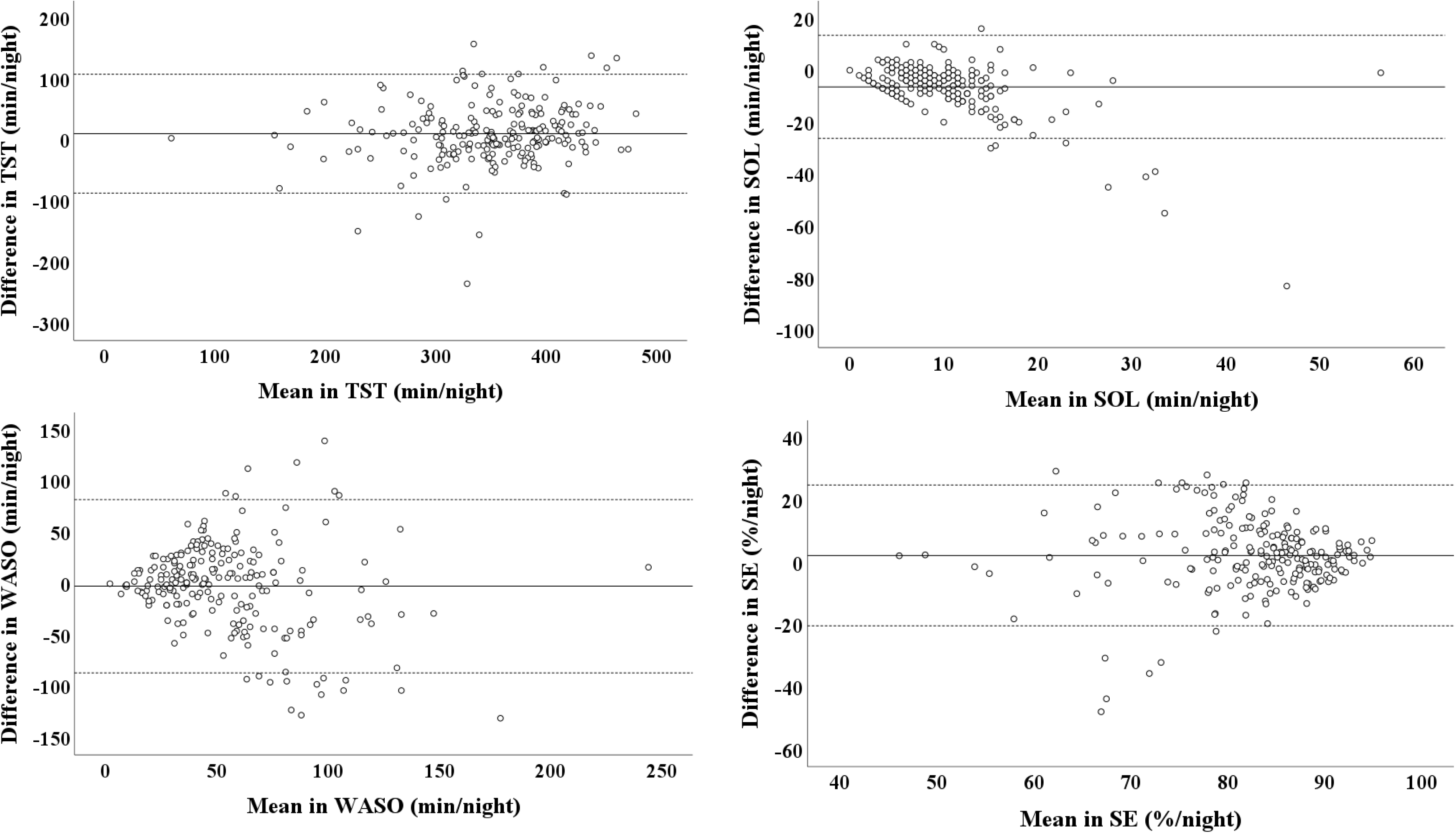
Bland–Altman plots comparing sleep parameters measured by wrist actigraphy (ActiGraph wGT3X-BT+) and portable EEG. The x-axis represents the mean of the two methods, and the y-axis represents the paired difference (actigraphy minus portable EEG). Solid lines indicate the mean difference, and dashed lines indicate the 95% limits of agreement. TST: Total Sleep Time, SOL: Sleep Onset Latency, WASO: Wake After Sleep Onset.

### 3.4 Magnitude of disagreement

The magnitude of disagreement varied across sleep parameters. For TST, the MAE and MAPE were 35.9 min and 11.1%, respectively. For SOL, the MAE and MAPE were 7.5 min and 59.1%; for WASO, 31.1 min and 83.6%; and for sleep efficiency, 8.45 percentage points and 11.1%.

### 3.5 Reliability between devices

ICCs showed parameter-specific variability in agreement (Table 3). Single-measure ICCs ranged from 0.227 to 0.728, whereas average-measure ICCs ranged from 0.370 to 0.843.

**Figure 4.**
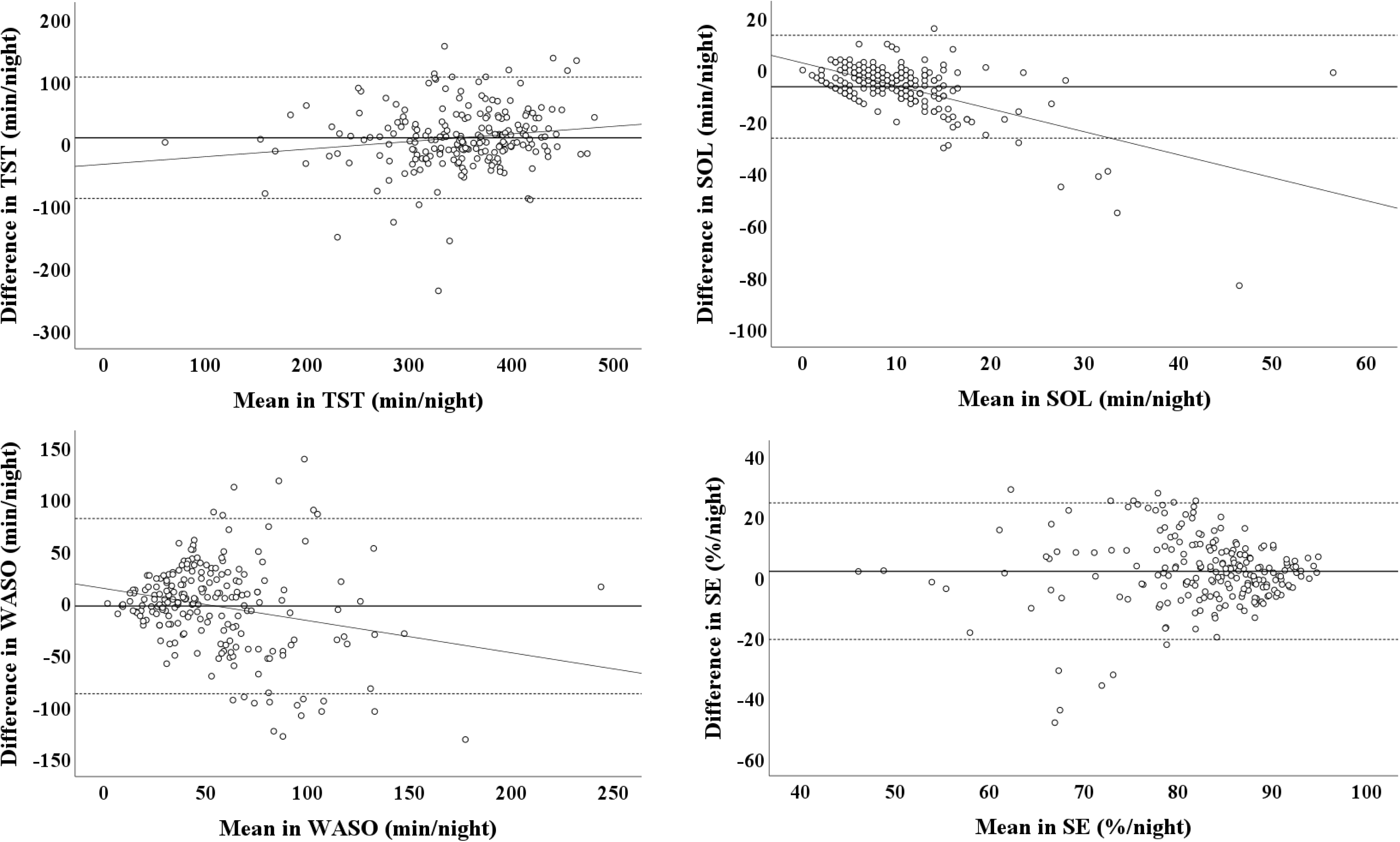
Bland–Altman plots comparing sleep parameters measured by wrist actigraphy (ActiGraph wGT3X-BT+) and portable EEG with fitted regression lines to assess proportional bias. The x-axis represents the mean of the two methods, and the y-axis represents the paired difference (actigraphy minus portable EEG). Solid lines indicate the mean difference, dashed lines indicate the 95% limits of agreement, and sloped lines indicate the fitted regression line (proportional bias). TST: total sleep time; SOL: sleep onset latency; WASO: wake after sleep onset.

**Table 3.**
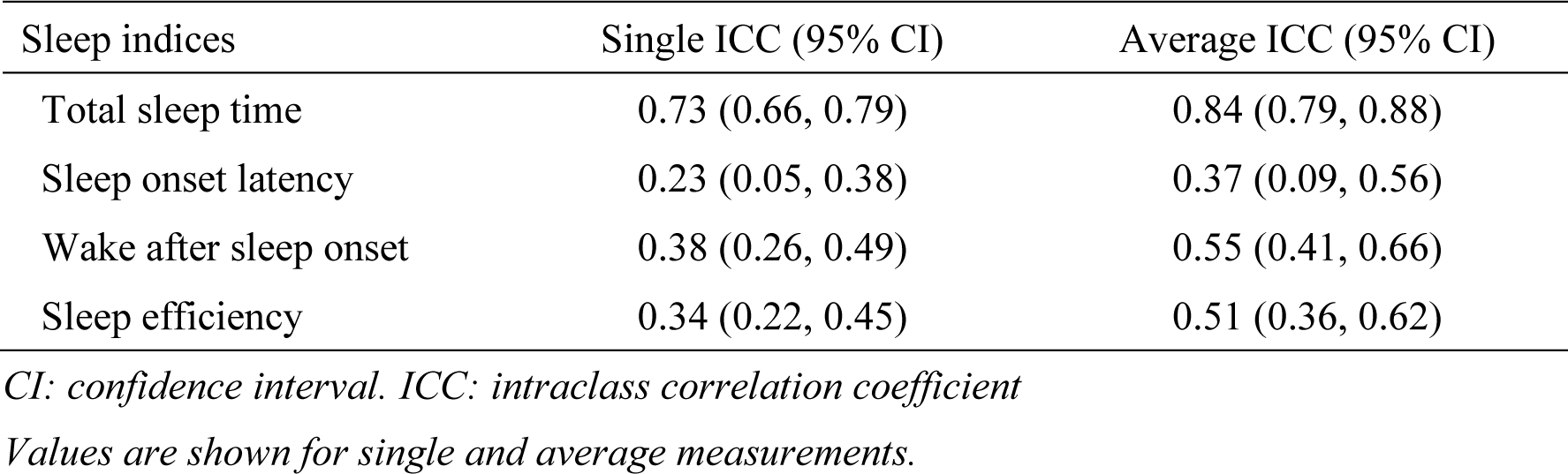
ICCs between actigraphy and portable EEG devices.

## 4. Discussion

In this validation study of community-dwelling older adults without major sleep or psychiatric disorders, wrist AG showed acceptable agreement with in-home portable EEG for average TST but clearly limited accuracy for SOL and WASO. AG systematically overestimated TST and sleep efficiency and underestimated SOL and WASO, with particularly large discrepancies at the single-night level. Averaging across multiple nights improved agreement for TST and, to a lesser extent, for WASO; however, substantial disagreement remained for SOL and WASO. Collectively, these findings support the use of wrist AG primarily for estimating habitual sleep duration in epidemiological research rather than for precise assessment of sleep initiation or nocturnal wakefulness in clinical practice.

### Comparison with previous studies

Previous studies in older adults have reported that actigraphy overestimates TST by 13–89 min and underestimates SOL by 3–13 min and WASO by 7–75 min relative to PSG- or EEG-based measures [37–39]. Our findings are broadly consistent with these reports, although the bias for TST in our sample was comparatively small. This likely reflects our exclusion of individuals with suspected sleep disorders, dementia, or severe depressive symptoms, thereby restricting the sample to older adults with relatively stable sleep patterns—an interpretation consistent with prior evidence that actigraphy can estimate TST with reasonable accuracy in individuals without sleep disorders [40]. Importantly, we observed these trends under free-living conditions using in-home portable EEG, strengthening the relevance of our findings to population-based sleep research and preventive interventions targeting older adults.

### Interpretation by agreement metrics (LoA, MAE/MAPE, ICCs)

Interpreting results across complementary agreement indices clarified parameter-specific performance. BA analyses demonstrated systematic differences between AG and portable EEG, and the LoA indicated that substantial uncertainty remained for SOL and WASO even when averaging across nights. Error-based indices supported this pattern: a MAPE of 10–15% has been suggested as acceptable in device-validation research [41], and TST met this criterion (MAPE = 11.1%), whereas SOL and WASO showed large relative errors. Notably, despite acceptable MAPE for TST, the MAE was approximately 36 min, indicating that discrepancies may still be meaningful for individual-level interpretation, particularly among those with very short or very long sleep durations. ICCs improved when values were averaged across multiple nights, particularly for TST and to a lesser extent for WASO, but agreement for SOL remained low to moderate. Together, these findings indicate that multi-night averaging improves stability but does not fully overcome the fundamental limitations of movement-based sleep inference for SOL and WASO. Because AG infers sleep–wake states from movement rather than directly measuring brain activity, subtle wakefulness and low-movement awakenings may be misclassified as sleep, likely contributing to the limited agreement for SOL and WASO [12].

### Implications

From a practical standpoint, wrist AG can reasonably serve as a primary tool for estimating habitual TST in large cohort studies, population-based lifestyle interventions, and community screening programs where group-level estimates and changes in average sleep duration are of primary interest. In such settings, AG offers scalable and low-burden monitoring when portable EEG or PSG is infeasible, enabling wider coverage at lower cost with an acceptable loss of precision. By contrast, when SOL or WASO constitutes a key clinical or trial outcome—such as in diagnostic evaluations for insomnia, titration or withdrawal of hypnotic medications, or clinical trials targeting sleep fragmentation—portable EEG or PSG should remain the primary assessment modality, with AG best positioned as an adjunct tool to capture broader sleep–wake patterns rather than to guide individual diagnostic or treatment decisions. This caution is consistent with previous reports highlighting actigraphy’s limitations in detecting brief wakefulness episodes [42,43].

### Limitations and future directions

Several limitations should be noted. First, the sample size was modest (49 participants), and individuals with suspected sleep disorders or cognitive decline were excluded, limiting generalizability to older adults with more disturbed sleep. Second, wearing both portable EEG and AG for several consecutive nights may have altered habitual sleep; however, this burden remains considerably lower than that associated with in-laboratory PSG. Third, AG scoring relied on a single sleep–wake algorithm (Cole–Kripke in ActiLife), and generalizability to other algorithms (e.g., Sadeh) or proprietary approaches remains uncertain. Future research should include participants with a broader range of sleep characteristics, examine algorithm-dependent differences, and test newer approaches—including machine-learning–based methods—to improve estimation of SOL and WASO and to narrow the LoA under free-living conditions. Consistent with evidence that actigraphy may not adequately capture the severity of sleep disturbance in older adults with severe insomnia [44], algorithm refinement will be essential to improve SOL/WASO estimation and to enhance clinical interpretability.

## 5. Conclusion

AG showed fair-to-good agreement with a portable EEG device for estimating TST and WASO in older adults when measurements were averaged over multiple nights, as indicated by ICCs and Bland–Altman analyses. This level of agreement supports the use of AG for assessing these sleep indices in large-scale epidemiological studies. However, bias remained substantial for SOL and sleep efficiency, and agreement for these indices was lower, suggesting that AG-derived estimates of SOL and sleep efficiency should be interpreted with caution at the individual level. Further refinement of AG algorithms that better capture the sleep characteristics of older adults, together with additional validation studies using different scoring algorithms, will be important for improving the clinical utility of AG.

## Glossary

AG: actigraphy
EEG: electroencephalography
ICC: intraclass correlation coefficient
LoA: limits of agreement
MAE: mean absolute error
MAPE: mean absolute percentage error
NREM: non–rapid eye movement sleep
PSG: polysomnography
REM: rapid eye movement sleep
SOL: sleep onset latency
TIB: time in bed
TST: total sleep time
WASO: wake after sleep onset

## Acknowledgements

We would like to thank Editage for editing and reviewing this manuscript for the English language.

## CRediT authorship contribution statement

Naoki Deguchi: Conceptualization, Formal analysis, Investigation, Methodology, Writing – original draft. Sho Hatanaka: Formal analysis, Writing – review & editing. Kaori Daimaru: Methodology, Project administration, Writing – review & editing. Kazushi Maruo: Formal analysis and Writing – review & editing. Hiroyuki Sasai: Conceptualization, Funding acquisition, Methodology, Supervision, Writing – review & editing.

## Data Availability

The datasets generated and/or analyzed during the current study are available from the corresponding author upon reasonable request.

## Declaration of generative AI and AI-assisted technologies in the manuscript preparation process

During the preparation of this manuscript, the authors used ChatGPT (OpenAI) to assist with language editing and improving clarity. After using this tool, the authors reviewed and edited the content as needed and take full responsibility for the content of the published article.

## Ethics approval statement

This study was approved by the Ethics Committee of Tokyo Metropolitan Institute for Geriatrics and Gerontology (approval number: R022-099).

## Patient consent statement

Written informed consent was obtained from all participants prior to their inclusion in the study.

## Permission to reproduce material from other sources

Permission to reproduce the figure from S’UIMIN Inc.

